# Impact of Case and Control Selection on Training AI Screening of Cardiac Amyloidosis

**DOI:** 10.1101/2023.03.30.23287941

**Authors:** Amey Vrudhula, Lily Stern, Paul C Cheng, Piero Ricchiuto, Chathuri Daluwatte, Ronald Witteles, Jignesh Patel, David Ouyang

**Author notes:** **Address for correspondence:** David Ouyang M.D., Staff Physician and Assistant Professor / Smidt Heart Institute, Cedars-Sinai Medical Center. **Funding:** None. **Disclosures:** PR and CD are employees of Alexion Pharmaceuticals. AV, LS, PCC, RW, JP, and DO report no disclosures.

## Abstract

**Background:** Recent studies suggest that cardiac amyloidosis (CA) is significantly underdiagnosed. For rare diseases like CA, the optimal selection of cases and controls for artificial intelligence (AI) model training is unknown and can significantly impact model performance.

**Objectives:** This study evaluates the performance of ECG waveform-based AI models for CA screening and assesses impact of different criteria for defining cases and controls.

**Methods:** Models were trained using different criteria for defining cases and controls including amyloidosis by ICD 9/10 code, cardiac amyloidosis, patients seen in CA clinic). The models were then tested on test cohorts with identical selection criteria as well as population-prevalence cohorts.

**Results:** In matched held out test datasets, different model AUCs ranged from 0.660 to 0.898. However, these same algorithms exhibited variable generalizability when tested on a population cohort, with AUCs dropping to 0.467 to 0.880. More stringent case definitions during training result in higher AUCs on the similarly constructed test cohort; however representative population controls matched for age and sex resulted in the best population screening performance.

**Conclusions:** AUC in isolation is insufficient to evaluate the performance of a deep learning algorithm, and the evaluation in the most clinically meaningful population is key. Models designed for disease screening are best with matched population controls and performed similarly irrespective of case definitions.

## Introduction

Cardiac amyloidosis (CA), an underdiagnosed disease driven by myocardial deposition of misfolded amyloid protein, is a progressive condition responsible for substantial morbidity and mortality.^1–4^ While early epidemiological data suggested a very low prevalence, autopsy series have found transthyretin amyloid (ATTR) deposits in 20-40% of octogenarians.^5, 6^ CA has been reported to be the etiology in up to 13% of patients with heart failure with preserved ejection fraction (HFpEF), and ATTR CA is present in approximately 16% of patients with severe calcific aortic stenosis undergoing transcatheter aortic valve replacement.^7–9^ Newer data with more sensitive imaging suggest the prevalence might be as high as 1-2% of the general population.^10^ Regional disparities in CA diagnosis also exist and particularly impact black Americans, who are disproportionately affected by the hereditary form of ATTR but are significantly undiagnosed.^11^

CA is challenging to diagnose because the disease is often indolent, and the symptoms are often similar to those of more common cardiac conditions. The disease is frequently not recognized until it has progressed to advanced stages, at which point treatment options are more limited. Timely diagnosis is vital, as new highly effective targeted therapies have been introduced for both light chain (AL) and ATTR CA, and these treatments display the greatest benefit when started early in the disease course.^12–14^ Previous work has demonstrated artificial intelligence’s (AI) ability to precisely phenotype diseases and characterize subtle cardiac physiology.^15–19^ Deep learning models have therefore been proposed to screen for CA using a variety of different forms of input data.^20–23^ While such models have demonstrated strong performance, given the scarcity and underdiagnosis of CA, these models are often trained on limited datasets without external validation, and further investigation into how cohort selection influences model performance is warranted.^22–25^

In this study, we sought to evaluate the impact of case and control definitions in the training of an AI to identify CA. We chose electrocardiogram (ECG) waveforms for model input as ECGs are inexpensive, non-invasive, widely available, and frequently obtained during routine visits. A variety of selection criteria as well as different ways to balance characteristics between cases and controls in the training dataset were used to evaluate the impact of training design choices on AI model performance. By maintaining the same AI model architecture, type of input data, and site across the experiments, we sought to evaluate whether more or less stringent case and control definitions would impact population level screening performance.

## Methods

### Data sources and study population

The study included ECGs from patients receiving care at Cedars-Sinai Medical Center between 2005 and 2022. The data was split 80% for training/10% for internal validation/10% for testing on a patient level prior to model development such that all models developed, irrespective of inclusion and exclusion criteria, were trained on data from the train split and were evaluated on the held-out test split. All training cohorts were matched 1:10 on cases and controls coming from the training split. Models were evaluated on ECGs from the held-out test split, with cohorts matched cohort selection criteria as well as the entire test split (population prevalence).

ECG waveform data, acquired at a sampling rate of 500 Hz, were extracted as 10 second, 12×5000 matrices of amplitude values. ECGs with missing leads were excluded from the study cohort. Associated clinical data for each patient, including demographic and clinical characteristics (e.g., age, gender, BMI, cardiovascular disease), were obtained from the electronic health record (EHR). Disease diagnoses were identified by International Classification of Diseases (ICD) 9th and 10th edition codes, which were also obtained from the EHR. The institutional review boards of Cedars-Sinai Medical Center and Stanford Healthcare approved the study protocol.

### AI Model Design and Training

A convolutional neural network for ECG interpretation was designed to detect the presence of cardiac amyloidosis. The model architecture is similar to those previously described to evaluate post-operative outcomes and screen for chronic kidney disease.^26, 27^ The model was trained using the PyTorch Lightning deep learning framework to predict outcomes with the input of one 12-lead ECG. If the same patient had multiple ECGs, each ECG was considered an independent example during training. Models were initialized with random weights and trained using a binary cross entropy loss function for up to 100 epochs with an ADAM optimizer and an initial learning rate of 1e-2. Early stopping was performed based on the validation dataset’s area under the receiver operating curve (ROC). The best model was determined based on population level screening performance in identifying cases in the hold-out test cohort. This model was used for downstream analysis in Table 3 and Table 4.

#### Case and Control Definitions and Test Populations

Three progressively more selective definitions of amyloid were evaluated to understand the effect of case definition on model performance: The broadest case definition used were diagnosis of amyloidosis by ICD 9/10 code (n = 990) The second case definition was for CA, defined by a subset of patients from the first cohort but also having evidence of cardiac involvement (n = 686). Cardiac involvement was defined as having an abnormal IVS measurement, brain natriuretic peptide (BNP), or troponin within 180 days of ECG. The third case cohort were patients seen in cardiac amyloid clinic (n = 168) with documented diagnosis by biopsy, Technetium-99m pyrophosphate (PYP) scintigraphy, or laboratory studies (serum free light chain, as well as serum and urine immunofixation) for monoclonal protein assessment.

Different populations of non-amyloid patients were chosen as controls. Control cohorts chosen for comparison include all non-amyloid patients, non-amyloid patients with left ventricular hypertrophy, non-amyloid patients with heart failure, and non-amyloid patients with heart failure with reduced ejection fraction (HFrEF). In various experimental setups, cases and controls were matched on different combinations of age, sex, wall thickness, and QRS amplitude to understand how these variables affected model performance. Wall thickness measurements were obtained from the closest echocardiogram within 180 days of the ECG. Case and controls ratios were always 1:10, except for HFrEF, where a ratio of 1:4.5 was used as HFrEF cases were uncommon in the control set.

### Statistical Analysis

A hold-out test dataset which was never seen during model training was used to assess model performance. Model performance was assessed by testing on three different test cohorts: 1.) a cohort mirroring the training and internal validation criteria in definitions and ratios of case and controls, 2.) the general population with cases defined as patients with an ICD 9/10 codes for amyloidosis (Supplementary Table 1), 3.) the general population with cases defined as amyloid clinic patients. The best model was determined based on population level screening performance in identifying cases in the hold-out test cohort. Model performance in identifying CA was assessed via area under receiver-operating curve (AUC). Two-sided 95% confidence intervals were computed using 10,000 bootstrapped samples for each metric. Statistical analysis was performed in R and Python.

## Results

### Population Characteristics

Our primary cohort consisted of 1,344,372 ECGs from 341,989 patients at Cedars-Sinai Medical Center. Amyloidosis cases comprised 10,042 ECGs across 990 patients, with cardiac amyloidosis representing 7,507 ECGs across 686 patients, and clinic patients represented by 2,256 ECGs from 168 patients. Demographics, comorbidities, and ECG characteristics are detailed in Table 1. Compared to controls, amyloid cases had a higher proportion of males, a higher proportion of black individuals, and the average age was older.

**Table 1:** Case and Control Demographics

|  | Amyloid by ICD9/10 | Cardiac Amyloidosis | Curated Cardiac Amyloidosis Clinic | Controls |
| --- | --- | --- | --- | --- |
| Number of ECGs | 10,042 | 7,507 | 2,256 | 1,334,330 |
| Number of Patients | 990 | 686 | 168 | 340,999 |
| Male Sex | 70.17% (7046) | 72.97% (5478) | 85.55% (1930) | 53.78% (715,999) |
| Age (years) | 70.62 (11.92) | 70.44 (11.94) | 70.26 (9.68) | 63.83 (18.87) |
| Black Race | 23.76% (2380) | 25.35% (1902) | 23.96 % (540) | 16.47% (218,049) |
| Hypertension | 48.46% (4866) | 50.66 % (3803) | 45.26% (1021) | 31.46% (419,750) |
| Diabetes Mellitus | 23.02% (2312) | 24.51% (1840) | 28.65% (632) | 15.57% (207,725) |
| Coronary Artery Disease | 45.32% (4552) | 48.39% (3633) | 43.34% (956) | 24.51% (327,050) |
| Heart Failure/Cardiomyopathy | 36.99% (3,715) | 27.00% (2,027) | 16.18% (357) | 21.80% (290,898) |
| IVS (cm)* | 1.343 (0.396) | 1.385 (0.392) | 1.433 (0.410) | 1.133 (0.491) |
| LVPW (cm)* | 1.318 (0.363) | 1.358 (0.359) | 1.376 (0.372) | 1.106 (1.028) |
| IVS or LVPW > 1.2 cm* | 61.74% (4875) | 68.27% (4546) | 66.43% (1320) | 34.98% (239,161) |
| ECG Amplitude (mV) | .633 (.295) | .632 (.310) | .614 (.390) | .695 (.618) |
| Heart Rate (bpm) | 81.63 (21.83) | 83.65 (22.14) | 86.25 (21.51) | 80.89 (21.90) |
| **<u>Abnormal ECG</u>** | 63.92% (6418) | 68.03% (5107) | 61.92% (1397) | 51.86% (691,990) |
| Bundle Branch Block | 17.71% (1779) | 18.90% (1419) | 19.24% (434) | 12.18% (162,481) |
| Ischemia | 13.02% (1307) | 14.80% (1111) | 11.44% (258) | 10.06% (134,230) |
| Infarct | 29.74% (2986) | 32.86% (2467) | 29.48% (665) | 17.79% (237,326) |
| Right Axis Deviation | 1.30 % (131) | 1.48% (111) | 1.51% (34) | 0.69% (9243) |
| Left Axis Deviation | 17.21% (1728) | 18.25% (1370) | 17.33% (391) | 9.393% (125334) |

Abbreviations: ECG – Electrocardiogram; IVS – Interventricular Septum, LVPW – Left Ventricular Posterior Wall; bpm – beats per minute; mV – millivolts;
\* Averages, Counts, and Percentages for these variables are based on the total number of patients with nearest laboratory testing or echocardiogram study within 180 days

### Model Performance in Defined Cohort and Screening Population

After optimizing case and control selection criteria and matching, our final model identified CA with an AUC of 0.820 (95% CI: 0.782 - 0.857) in the general population and an AUC of 0.744 (95% CI 0.721 - 0.767) in the matched held-out test set. The best model used cases with cardiac amyloidosis and controls matched by age and sex for model training, with both cardiac amyloidosis by ICD9/10 code definition and seen in CA clinics achieved similar performance (Table 2, Central Illustration).

**Table 2:** Model Performance in Defined Cohort and Screening Population

|  | <u>Cases Criteria</u> | <u>Controls Criteria</u> | Matching | Matched Cohort AUC | Population AUC (Cases by ICD9/10) | Population AUC (Cases by Clinic List) |
| --- | --- | --- | --- | --- | --- | --- |
| Varying Case Definition | Amyloid by ICD9/10 | Non-amyloid | None | 0.705<br>(0.679 - 0.730) | 0.702<br>(0.677 - 0.727) | 0.844<br>(0.803 - 0.88) |
|  | Amyloid by ICD9/10 w/<br>Cardiac involvement | Non-amyloid | None | 0.750<br>(0.726 - 0.773) | 0.728<br>(0.706- 0.751) | 0.866<br>(0.830 - 0.898) |
|  | Curated Amyloid Clinic List | Non-amyloid | None | 0.880<br>(0.844 - 0.912) | 0.720<br>(0.695 - 0.744) | 0.880<br>(0.844- 0.911) |
| Varying Control Definition | Amyloid by ICD9/10 | Non-amyloid<br>HFrEF | None | 0.767<br>(0.745- 0.789) | 0.467<br>(0.443 - 0.491) | 0.426<br>(0.378- 0.476) |
|  | Amyloid by ICD9/10 | Non-amyloid Heart<br>Failure | None | 0.650<br>(0.625 - 0.674) | 0.517<br>(0.491- 0.544) | 0.545<br>(0.494 - 0.606) |
|  | Amyloid by ICD9/10 | Non-amyloid LVH | None | 0.660<br>(0.642 - 0.736) | 0.570<br>(0.545 - 0.596) | 0.690<br>(0.642 - 0.736) |
| Varying Case to Control Matching | Amyloid by ICD9/10 | Non-amyloid | Age and Sex | 0.682<br>(0.656 - 0.708) | 0.698<br>(0.674 - 0.722) | 0.861<br>(0.828 - 0.892) |
|  | Amyloid by ICD9/10 | Non-amyloid | Age, Sex, and<br>Wall Thickness | 0.662<br>(0.633 - 0.691) | 0.659<br>(0.633 - 0.685) | 0.819<br>(0.778 - 0.856) |
|  | Amyloid by ICD9/10 | Non-amyloid | Age, Sex, and<br>QRS amplitude | 0.677<br>(0.633 - 0.691) | 0.636<br>(0.609 - 0.662) | 0.775<br>(0.730-0.818) |
| Best Models | Amyloid by ICD9/10 w/<br>Cardiac Involvement | Non-amyloid | Age and Sex | 0.744<br>(0.721 - 0.767) | 0.733<br>(0.711 - 0.754) | 0.820<br>(0.782 - 0.857) |
|  | Curated Amyloid Clinic List | Non-amyloid | Age and Sex | 0.898 | 0.714 | 0.898 |
|  |  |  |  | (0.868 - 0.924) | (0.690 - 0.738) | (0.870- 0.923) |

### Performance of Varying Case and Controls

When varying case definition, we found that stricter case inclusion criteria resulted in increased AUC when tested on the matched held-out test set (AUC increasing from 0.705 to 0.880 with increased stringency), however the improved performance did not generalize when tested on the population cohort (AUCs ranging from 0.702 to 0.728 with overlapping confidence intervals). Models in which cases and controls were matched for QRS amplitude performed the poorest of these matching combinations when tested on population level cohorts for both cardiac amyloidosis ICD definitions and clinic cases definitions.

Models trained against controls that were phenotypically most distinct from amyloid resulted in the highest AUC in the matched test cohorts (for example, with HFrEF controls, the AUC was 0.767 (95% CI: 0.745-0.789), while controls with greater overlap with amyloid demonstrated lower AUCs in the matched cohorts (Non-amyloid LVH controls resulted in an AUC of 0.660 (95% CI: 0.642 - 0.736). However, these results on matched cohorts did not correlate with population level AUCs (Figure 1), particularly in models in which the inclusion criteria for controls during testing were significantly different from the general population. Models with LVH controls generalized better to the population test sets (AUC 0.570) compared to models trained with HFrEF (AUC 0.467) or heart failure (AUC 0.517) controls.

**Figure 1.**
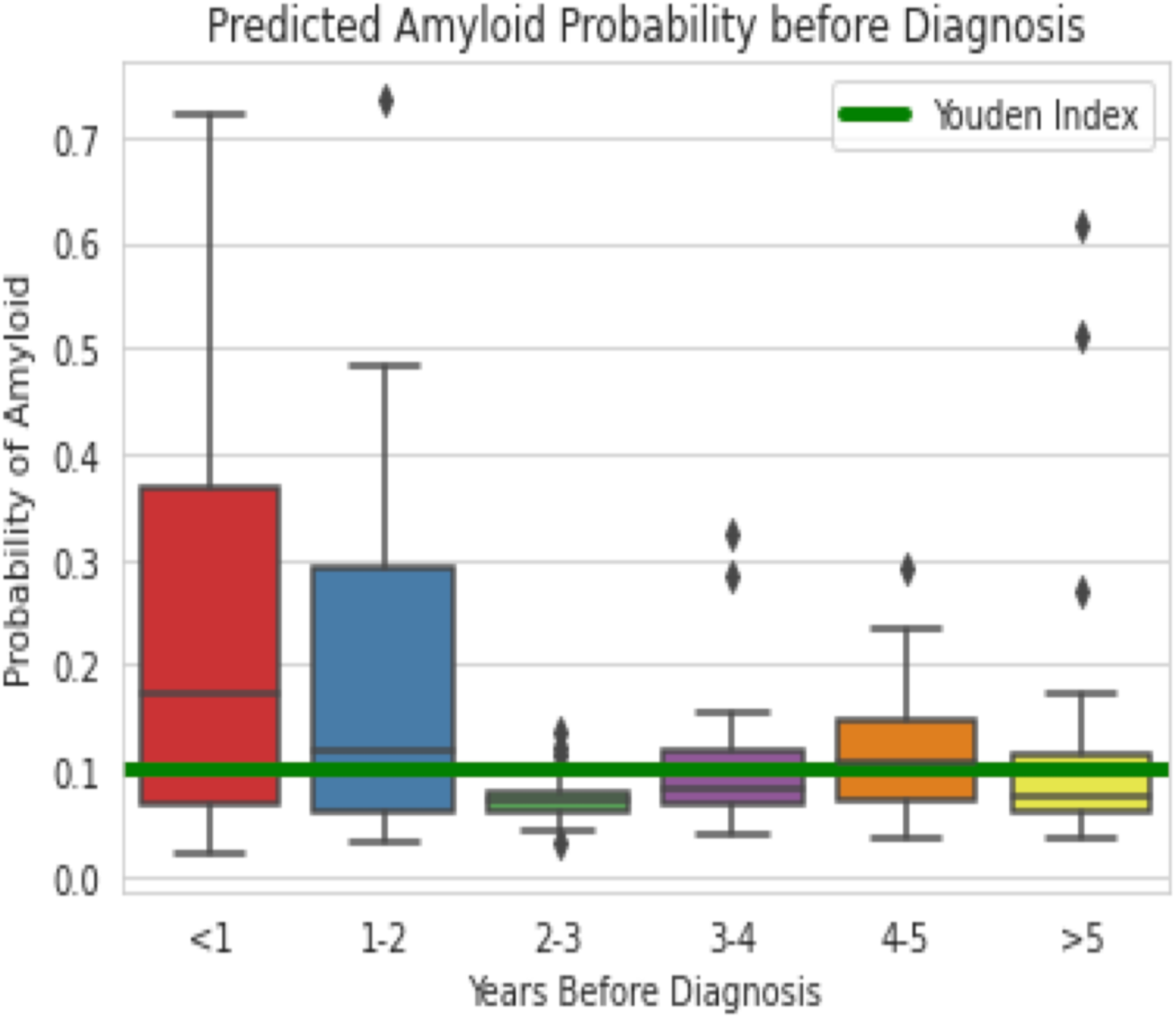
Predicted Amyloid Probability before diagnosis: Probability of amyloid (y-axis) is shown here for ECGs taken before diagnosis to assess if the model can detect amyloidosis before date of clinical diagnosis.

For all models, the choice of case definition in the held-out population test split significantly impacted AUC, as the AUC was consistently higher when cases were defined by clinic adjudication rather than by ICD9/10 definition (mean difference of 0.116 (0.068) across models). For downstream analyses, we chose the ICD9/10 definition of cases for evaluation of sensitivity/specificity as this difference in performance was likely due to later and more obvious phenotypes being seen in the clinic cohort; as we envision the use of such an algorithm for screening, the goal is to identify earlier cases.

### Secondary Analyses

We sought to understand the utility of the model as a screening tool by measuring sensitivity, specificity, and predictive values at various screening thresholds (Table 3). At the Youden Index, the model showed a sensitivity of 0.609 (95% CI: 0.569 - 0.648) and specificity of 0.718 (95% CI: 0.714 - 0.721). PPV was 0.018 (95% CI: 0.016 - 0.020) while NPV was 0.995 (95% CI: 0.995 - 0.996). With a chosen specificity of 0.974, an estimated additional 275 patients would be identified out of every 10,000 patients screened. Several populations are at higher risk for amyloidosis, and we examined model performance in these groups (Table 4). The AUC for males (0.756 (95% CI: 0.731 - 0.781)) and individuals greater than 60 years of age (0.739 (95% CI: 0.716 - 0.763)) were consistently high.

**Table 3:** Assessing Model Utility As a Screening Tool at Different Thresholds

| **Threshold** | **Sensitivity** | **Specificity** | **PPV** | **NPV** | **Positives per**<br>**10,000 screened** |
| --- | --- | --- | --- | --- | --- |
| **0.101**<br>**(Youden Index)** | 0.609<br>(0.569 - 0.648) | 0.718<br>(0.714 - 0.721) | 0.018<br>(0.016 - 0.020) | 0.995<br>(0.995 - 0.996) | 2,853<br>(2,818 - 2,887) |
| **0.25** | 0.212<br>(0.178 - 0.246) | 0.974<br>(0.973 - 0.975) | 0.064<br>(0.053 - 0.076) | 0.993<br>(0.993 - 0.994) | 275<br>(263 - 288) |
| **0.5** | 0.076<br>(0.056 - 0.100) | 0.997<br>(0.996 - 0.997) | 0.162<br>(0.120 - 0.207) | 0.992<br>(0.992 - 0.993) | 40<br>(35 - 45) |
| **0.7** | 0.023<br>(0.011 - 0.036) | 0.999<br>(0.999 - 1.000) | 0.241<br>(0.132 - 0.361) | 0.992<br>(0.991 - 0.992) | 8<br>(6 - 10) |

**Table 4:** Assessing Model Performance in Population Subsets

| Population Subset | N | Similar Cohort<br>AUC | N | Population AUC | Population<br>Sensitivity | Population<br>Specificity |
| --- | --- | --- | --- | --- | --- | --- |
| All EKGs | 10,648 | 0.744<br>(0.721 - 0.767) | 134,845 | 0.733<br>(0.712 - 0.755) | 0.609<br>(0.569 - 0.649) | 0.718<br>(0.714 - 0.721) |
| Male | 7,150 | 0.760<br>(0.733 - 0.787) | 71,476 | 0.756<br>(0.731 - 0.781) | 0.669<br>(0.622 - 0.716) | 0.691<br>(0.686 - 0.696) |
| Age ≥ 60 | 8,536 | 0.764<br>(0.735 - 0.784) | 84,648 | 0.739<br>(0.716 - 0.763) | 0.651<br>(0.606 - 0.695) | 0.686<br>(0.682 - 0.690) |
| African American Race | 1,801 | 0.664<br>(0.622 - 0.705) | 21,930 | 0.681<br>(0.645 - 0.715) | 0.491<br>(0.420 - 0.560) | .718<br>(0.709 - 0.726) |
| LVH Documented by<br>Echo | 899 | 0.581<br>(0.525 - 0.633) | 39,755 | 0.696<br>(0.668 - 0.723) | 0.674<br>(0.627 - 0.718) | 0.578<br>(0.571 - 0.585) |
| Normal Heart Rate | 7,767 | 0.741<br>(0.714 - 0.767) | 96,722 | 0.737<br>(0.712 - 0.761) | 0.600<br>(0.549 - 0.642) | 0.733<br>(0.729 - 0.737) |
| Heart Failure or<br>Cardiomyopathy | 3,146 | 0.748<br>(0.693 - 0.801) | 38,908 | 0.754<br>(0.710 - 0.796) | 0.633<br>(0.556 - 0.709) | 0.714<br>(0.708 - 0.720) |

*\*Sensitivity and specificity were calculated using the Youden Index as the threshold*

Given that identifying CA early is paramount to reaping the greatest benefit from new therapies, we sought to gauge the model’s ability to predict CA prior to diagnosis. Detailed model predictions of EKGs before clinical diagnosis date is shown in Figure 1. Overall, at the Youden index, the model detected disease in a significant proportion of EKGs at least 2 years before first diagnosis. With the strong relationship between QRS amplitude and LVH, EKGs carry information that can be leveraged to predict wall thickness. We show that deep learning can predict wall thickness with a mean absolute error of 2 mm (Figure 2). However, it should be noted that matching based on wall thickness measurements from echocardiograms or LVH (wall thickness > 1.2 cm) did not improve population screening performance of the model, and similarly wall thickness alone was insufficient to identify CA.

**Figure 2a.**
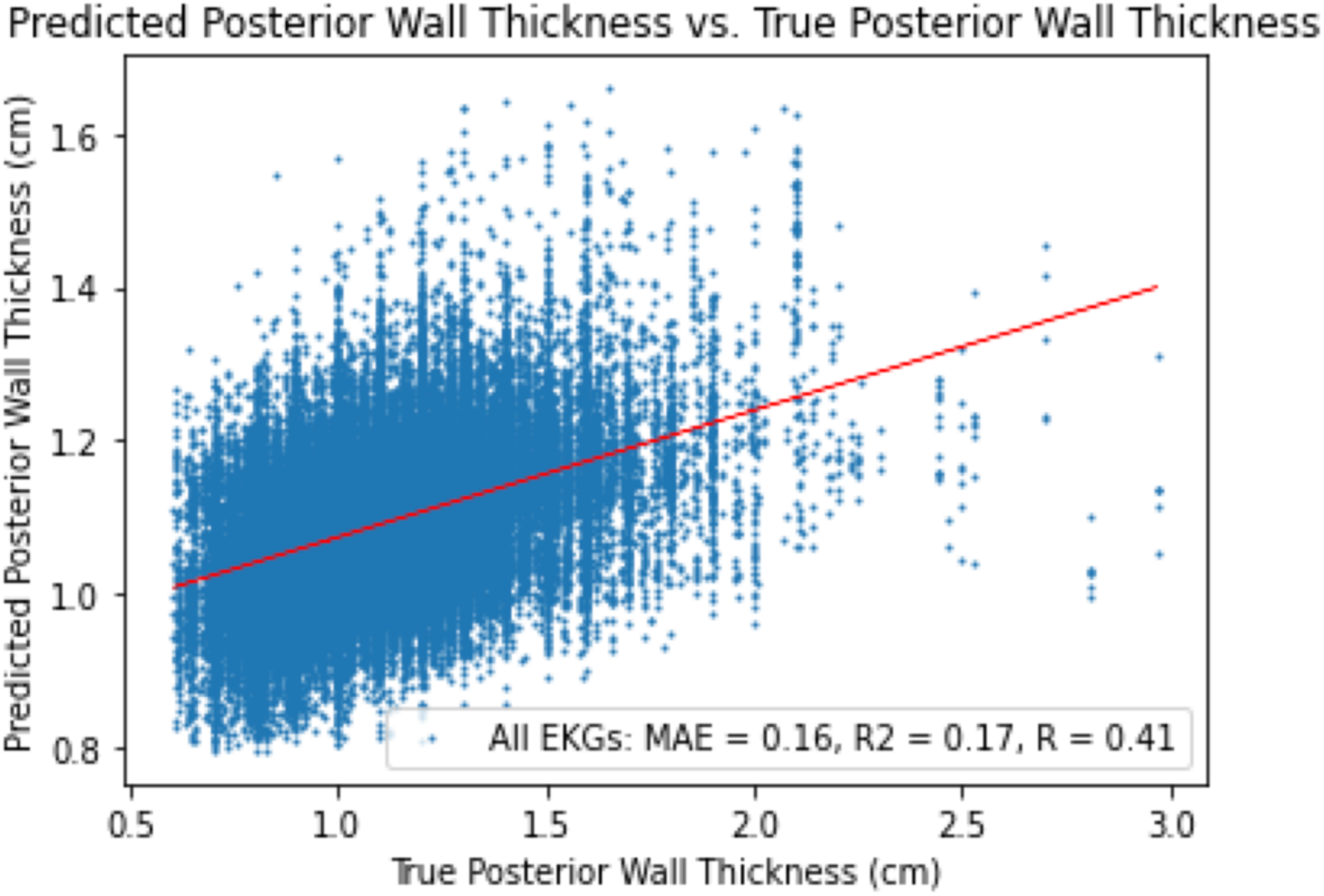
Predicted LVPW Thickness: Prediction of Left Ventricular Posterior Wall (LVPW) Thickness from ECGs using a deep learning model.

**Figure 2b.**
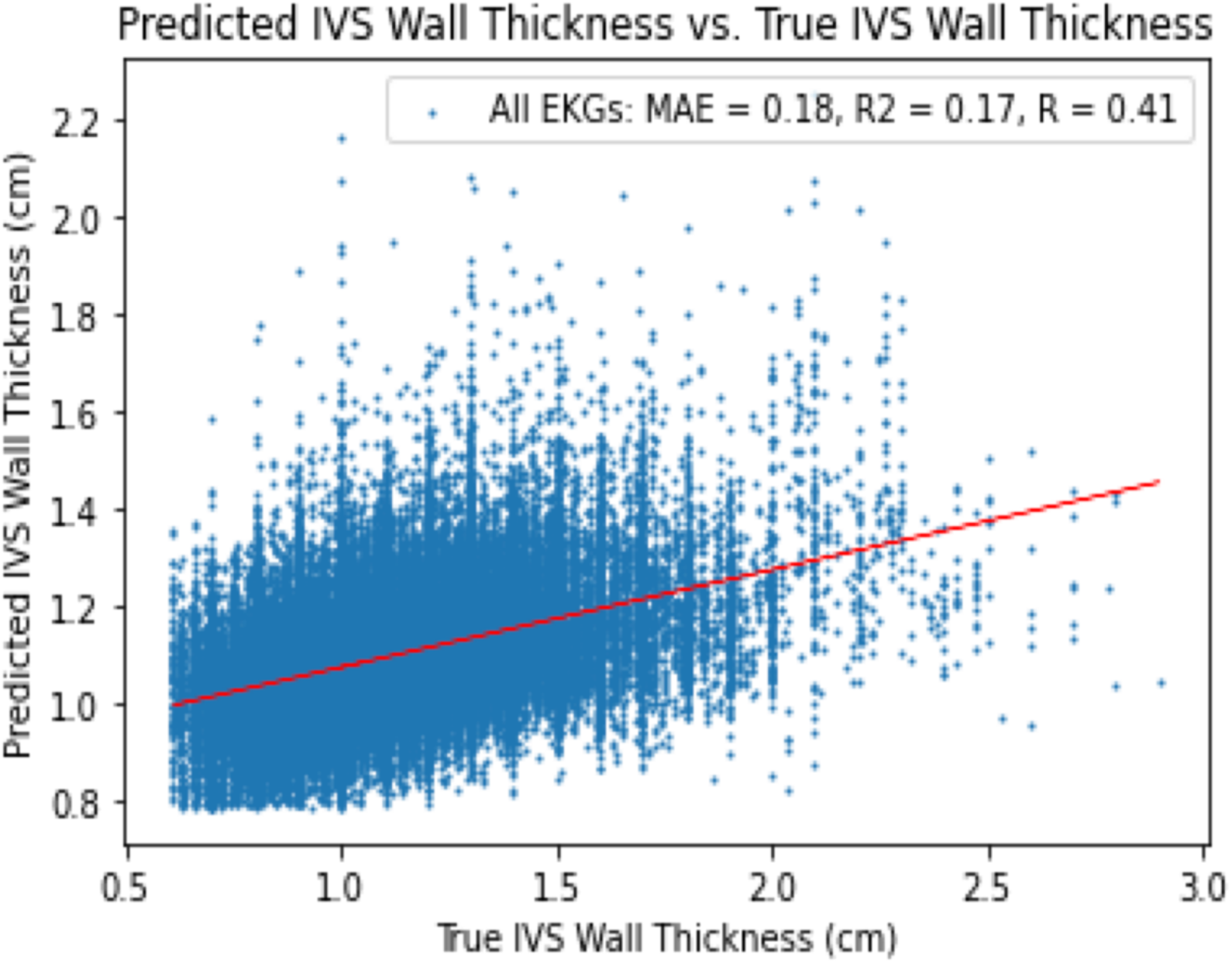
Predicted IVS Thickness: Prediction of interventricular septal (IVS) thickness from ECGs using a deep learning model.

## Discussion

In this study, we reaffirm prior analyses that cardiac amyloidosis can be identified in ECGs through AI evaluation.^22, 23^ Additionally, we also show that case and control selection influences model performance and generalizability, with more stringent inclusion criteria not always generalizing to the best model for population screening. We show that testing with clinic-derived lists of CA result in higher AUC, likely due the over-representation of later and more fulminant cases, however on a population level, models perform similarly in identifying patients with CA. AUC alone is an imperfect metric in assessing deep learning models, and the choice of cases and controls should be taken into account in training AI models.

Choosing the training cohorts of an AI model is similar to designing a case-control study. Seemingly small choices in selection criteria can result in large impacts on the generalizability and performance of the AI models, as AI models can identify shortcut variables and confounding influences and can therefore be biased towards more severe phenotypes. In the case of CA, clinic cohorts likely are enriched for more severe phenotypes, including patients who are being considered for advanced therapies, which might be easier to identify but not necessarily the optimal patients to identify via screening. These cases might not be representative of subtle early cases which one hopes to identify in disease screening and the model performance might be overestimated as more severe cases are easier to identify.

As CA is a disease historically been considered rare, training of AI models for identifying CA is limited by having few well-phenotyped cases for model training. Fortunately, we see in our experiments that potentially less well-curated lists result in similar performance to models trained on well characterized clinic lists. The ability to train models even on less curated patient lists may then open the door for institutions without specific amyloid clinics to refine or train models for screening among populations at risk. Equally important is the choice of controls and how cases and controls are matched during model training.

There are a few limitations of note. First, there are limitations to disease definitions by ICD9/10 codes. While we show similar results with our curated clinic cohort, many patients with amyloidosis by ICD9/10 codes do not have confirmatory testing available for review in the electronic health record. Second, our selection of cardiac involvement requires selected laboratory testing or echocardiographic assessment to be done within 180 days, which might bias towards more severe or obvious cases of CA. While this is similarly true for patients in amyloid clinic, this degree of label noise might both result in false positives and false negatives. Further work is required to bring these findings to the bedside. Importantly, given the underdiagnosis of amyloidosis, there are likely patients in the control cohort who have undiagnosed CA, limiting the potential model performance. Additionally, most models for screening of CA are restricted to a few centers, so validating this model at other centers is key to understanding the generalizability of these AI tools. Finally, a truly prospective study is necessary to gauge the true clinical impact of screening AI models.

## Conclusion

Cardiac amyloidosis is an underdiagnosed progressive disease with phenotypic heterogeneity, and care should be taken in understanding how AI models are trained to screen for CA. In this study, we found selection of cases and controls significantly impacts model performance on a general population, with even less well-phenotyped case definition being able to train AI models. AUC alone is insufficient to assess the generalizability of these AI models, and further external validation as well as prospective validation is needed to understand the utility of screening AI models.

## Data Availability

All data produced in the present study are available upon reasonable request to the authors.

## Abbreviations list

(CA): Cardiac Amyloidosis
(AI): Artificial Intelligence
(ECG): Electrocardiogram
(ROC): Receiver Operating Curve
(AUC): Area Under the Curve
(BNP): Brain Natriuretic Peptide
(ICD): International Classification of Diseases
(ATTR): transthyretin amyloid
(AL): light chain amyloid
(LVPW): Left Ventricular Posterior Wall
(IVS): Interventricular Septum
(EHR): Electronic Health Record

## Acknowledgements

*A*.*V. is a research fellow supported by the Sarnoff Cardiovascular Research Fellowship*.

## Figure Legend

**Central Illustration:**
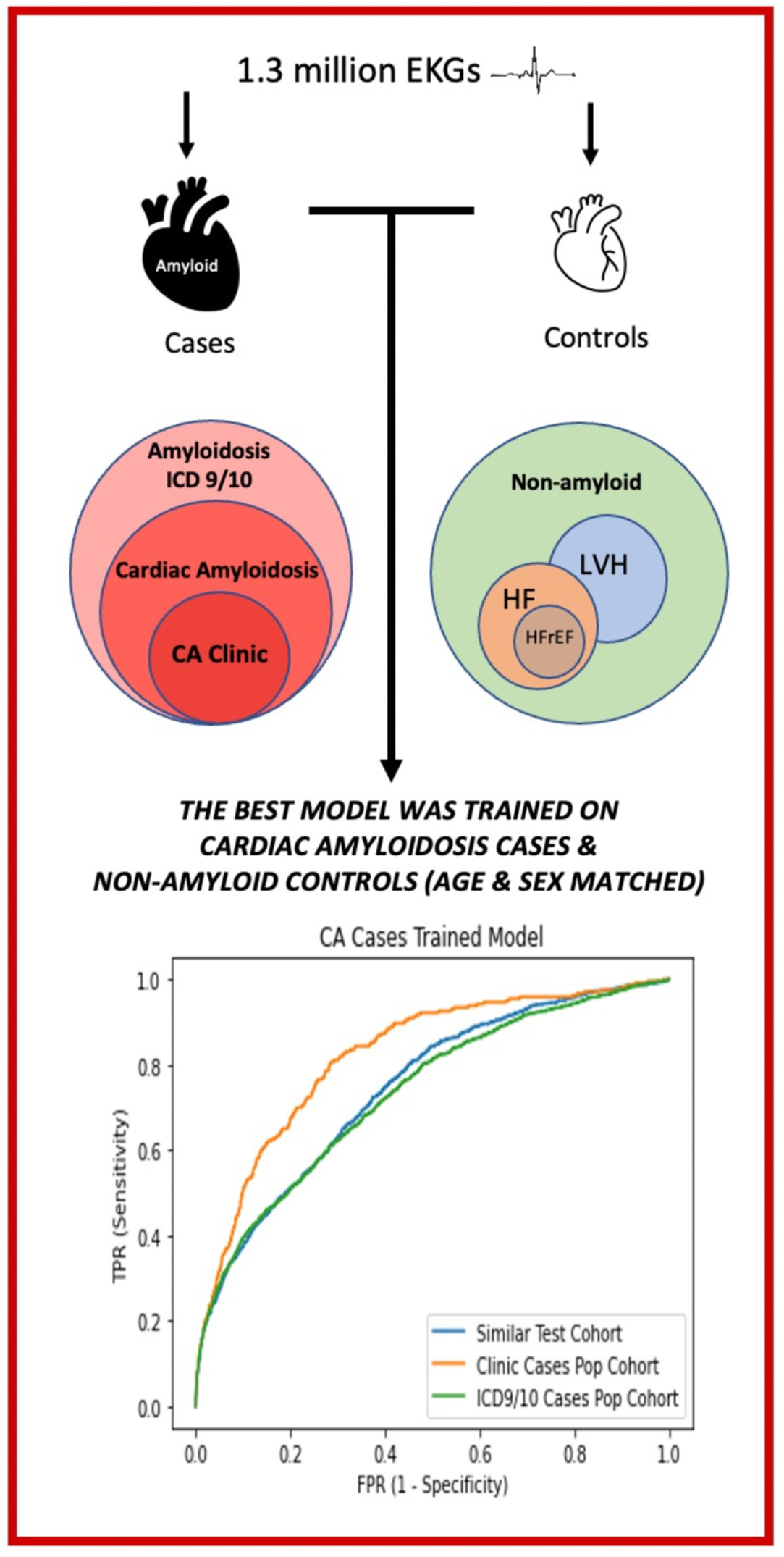
Best CA Detection Models Trained with CA cases and Age & Sex Matched Non-amyloid Controls: Model performance was evaluated while training with different case and control definitions. The best model trained on cardiac amyloidosis cases and non-amyloid controls matched for age and sex. These results show that models trained on less stringent case definitions perform just as well, if not better, on a population level than highly phenotyped cases. These results open the door for centers without dedicated amyloid clinics to train models that could potentially be used as population screening tools.

