## Supplemental Table 1 for "Impact of Case and Control Selection on Training AI Screening of Cardiac Amyloidosis"

*Supplemental Table 1 - ICD Codes*

| ***Code*** | ***Name*** | ***Code Type*** |
| --- | --- | --- |
| *277.3* | *Amyloidosis* | *ICD 9* |
| *277.30* | *Amyloidosis* | *ICD 9* |
| *277.39* | *Other amyloidosis* | *ICD 9* |
| *E85* | *Amyloidosis* | *ICD 10* |
| *E85.0* | *Non-neuropathic heredofamilial amyloidosis* | *ICD 10* |
| *E85.1* | *Neuropathic heredofamilial amyloidosis* | *ICD 10* |
| *E85.2* | *Heredofamilial amyloidosis* | *ICD 10* |
| *E85.4* | *Organ-limited amyloidosis* | *ICD 10* |
| *E85.8* | *Other-limited amyloidosis* | *ICD 10* |
| *E85.81* | *Light chain (AL) amyloidosis* | *ICD 10* |
| *E85.82* | *Wild-type transthyretin-related (ATTR) amyloidosis* | *ICD 10* |
| *E85.89* | *Other amyloidosis* | *ICD 10* |
| *E85.9* | *Amyloidosis* | *ICD 10* |
